# Integrative Proteomic and Metabolomic Signatures of Accelerated PhenoAge in the UK Biobank

**DOI:** 10.64898/2026.03.02.26347435

**Authors:** Kamila Bloch, João Pedro de Magalhães

## Abstract

Aging is accompanied by molecular changes across multiple biological systems that contribute to functional decline and increased disease risk, but the underlying mechanisms and inter-individual variation remain poorly understood. We investigated whether multi-omics integration can reveal coordinated molecular processes associated with accelerated PhenoAge, a clinical biomarker-based estimate of biological aging. Using UK Biobank data from ∼20,000 participants, we integrated plasma proteomics and metabolomics with PhenoAge. Individuals were stratified by extreme PhenoAge acceleration (*PhenoAgeAccel*) to enhance biological contrast. We applied a supervised multi-omics integration framework (DIABLO) to identify correlated molecular features that distinguish accelerated from slower PhenoAge. Proteomics alone provided strong discrimination between aging groups, whereas metabolomics showed weaker performance. Integrating both modalities did not substantially improve classification accuracy but revealed four multi-omics modules: immune-inflammatory, lipid-vascular, nutrient-metabolic, and HDL-apolipoprotein pathways. The dominant signature reflected immune and inflammatory activation with metabolic support, consistent with established aging processes, while additional components highlighted lipid transport, vascular signaling, and nutrient regulation. Nearly half of DIABLO-selected proteins overlapped curated aging and senescence databases, supporting relevance to aging. Together, these results show that multiomics integration enables the identification of coordinated proteomic-metabolomic axes associated with accelerated aging processes, enhancing biological interpretability beyond single-omics analyses.

## Introduction

Age is the strongest risk factor for many chronic diseases and mortality^1^. However, people of the same chronological age often differ in physiological function, disease burden, and survival. This heterogeneity suggests variation in the underlying processes of aging. In response, multiple biomarkers and “aging clocks” have been developed to quantify biological age and predict morbidity and mortality. Although the extent to which these measures capture biological aging itself is debated^2^, they provide insight into an individual’s health status and vulnerability^3^. Understanding the molecular processes associated with faster physiological decline and the associated increase in disease risk and mortality may therefore enable earlier risk detection and inform strategies aimed at extending healthspan, consistent with the goals of geromedicine^4^.

Although debated, a wide range of biological age estimators has been developed as proxies for biological age. Phenotypic measures, such as PhenoAge^5^, are derived from routine clinical biomarkers, while molecular clocks leverage epigenetic, transcriptomic, proteomic, or metabolomic data^6, 7^. Epigenetic clocks are the most established: first-generation models (e.g. Horvath^8^, Hannum^9^) were trained to predict chronological age, whereas second-generation models (e.g. DNAm PhenoAge^5^, GrimAge^10^) incorporate morbidity and mortality endpoints, directly reflecting underlying aging-related biology^6^. However, their biological interpretation remains challenging, due in part to sensitivity to cell-type composition and limited understanding of whether captured signals represent drivers or downstream consequences of aging^11, 12^.

Other omics layers may offer complementary insight. Transcriptomic studies have identified age-associated expression signatures^13, 14, 15^, while proteomic and metabolomic approaches have gained increasing attention. Plasma proteomics captures systemic dysregulation across immune, vascular, and metabolic pathways and has revealed organ-specific aging signatures^16, 17, 18, 19^. Large-scale proteomic aging models have shown robust associations with disease and mortality in diverse populations^19^. Metabolomic models also outperform chronological age for predicting morbidity and mortality in large cohorts^20, 21, 22, 23, 24^. Metabolomics contributes complementary information by integrating intrinsic metabolic processes with modifiable environmental and lifestyle exposures. Moreover, nuclear magnetic resonance (NMR) metabolomics, as implemented in UK Biobank^25^, is highly standardised, quantitative, and reproducible, with a relatively simple and low-cost workflow^26^, which facilitates scalability and potential clinical adoption. Together, these findings suggest that proteomic and metabolomic data capture biologically meaningful aspects of aging that may not be fully reflected by epigenetic clocks alone.

Despite these advances, it remains unclear which molecular processes distinguish individuals who age faster or slower than expected for their chronological age, and how different omics layers jointly contribute to this heterogeneity. Many approaches focus either on single-omics predictors or on improving predictive accuracy, often at the expense of biological interpretability. Here, we address this gap by integrating plasma proteomics and metabolomics with PhenoAge^5^, a clinically grounded measure of biological aging, in a large UK Biobank^25^ subcohort. We defined accelerated aging using *PhenoAgeAccel* and extreme phenotypes to enhance biological contrast rather than to focus on inferring population-level risk. We applied a multi-omics integration framework DIABLO, a supervised multivariate method within *mixOmics* package that simultaneously models multiple omics datasets to identify correlated features across data types^27^. Our aim was to identify multi-omics signatures that characterize accelerated aging and to assess whether integration provides biologically interpretable signatures beyond those captured by proteomics or metabolomics alone.

## Results

### Study population

After quality control (see Methods), the analytical sample included 18,632 UK Biobank participants with complete proteomic, metabolomic, and PhenoAge data. Participants were aged 39-70 years (mean = 56.7 ± 8.1), and 54.1% were female (**Table 1**). The age and sex distributions were similar to the whole UKB cohort. For multivariate analyses, individuals in the upper and lower 20% of the *PhenoAgeAccel* distribution were selected to enhance contrast and reduce overlap between borderline phenotypes.

**Table 1.** Study cohort characteristics.

| **Participants (n = 18,632)** |  |
| --- | --- |
| **Age (years)** |  |
| Mean (SD) | 56.69 (8.06) |
| Median (range) | 58.00 (39-70) |
| **Sex** |  |
| Female | 10,088 (54.1%) |
| Male | 8,544 (45.9%) |
| **PhenoAgeAccel (years)** |  |
| Mean (SD) | 0.05 (5.77) |

### Single-omics models (sPLS-DA)

Sparse Partial Least Squares Discriminant Analysis (sPLS-DA) was applied separately to proteomic and metabolomic data from individuals with extreme *PhenoAgeAccel* values, with chronological age and sex regressed out prior to model fitting.

The proteomic model achieved robust discrimination between accelerated and slower aging groups, with a balanced error rate (BER) of 0.15 across five components. Functional enrichment analyses revealed that the strongest signal was concentrated in the first two components, which were enriched for immune-inflammatory signaling, cytokine activity, cell adhesion, and growth factor pathways. Subsequent components showed minimal enrichment. In contrast, the metabolomic model required a larger number of components (eight) to achieve separation and demonstrated lower discriminative performance (BER = 0.233), suggesting weaker discriminatory capacity compared with proteomics. It is important to note that the number of proteins (n = 2,923) exceeded the number of metabolites (n = 248), which may in part have contributed to the stronger performance of the proteomic model. Together, these results indicate that plasma proteomics captures stronger discriminatory signal for *PhenoAgeAccel* than metabolomics when analysed in isolation.

### Integrative multi-omics model (DIABLO): performance and component-level separation

We next applied DIABLO to jointly model proteomic and metabolomic data from the same extreme *PhenoAgeAccel* subset. The final model included four components, selecting a sparse set of features from each omics block (30, 30, 20, and 20 proteins; 3, 4, 3, and 4 metabolites per component). Model performance was stable across internal cross-validation and the independent test set (Train BER = 0.176; Test BER = 0.168), indicating limited overfitting. As in single-omics analyses, discrimination was driven primarily by proteomic features (proteomics AUC = 0.91; metabolomics AUC = 0.77).

To quantify biologically meaningful group separation, we examined effect sizes of DIABLO component scores rather than relying solely on statistical significance (**Figure 1**). Component 1 showed strong separation between accelerated and slower aging groups, driven predominantly by proteomic features (Cohen’s d = −1.49), with supporting contribution from metabolites (d = −0.85). Component 2 reflected a shared proteomic-metabolomic axis with moderate to large effects in both blocks (proteomics d = 0.41; metabolomics d = 0.68). Component 3 showed weaker separation, and Component 4 had very small effect sizes despite statistical significance, suggesting limited biological relevance. These results indicate that the primary aging-related signal is concentrated in the first two DIABLO components.

**Figure 1.**
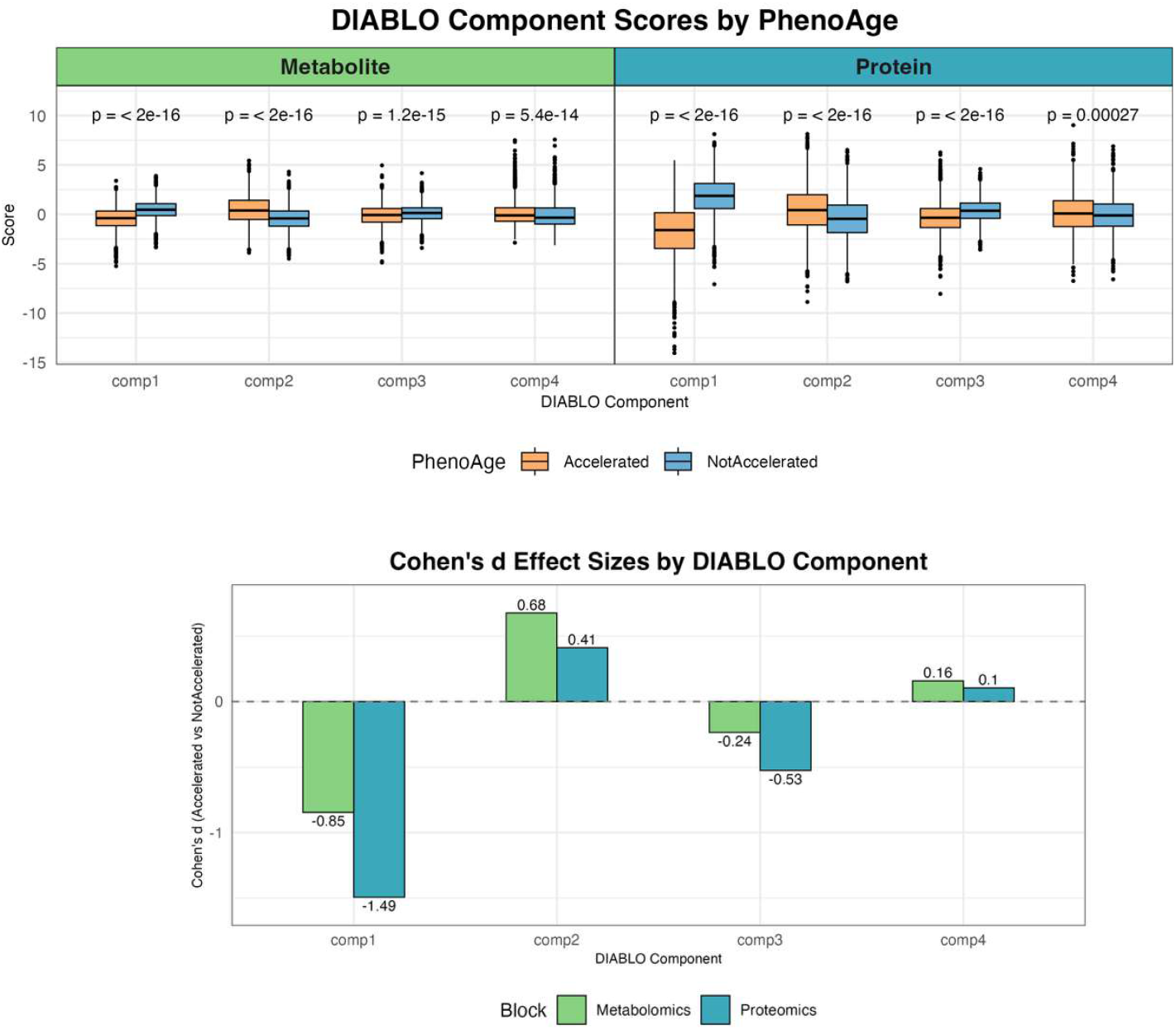
Group separation and effect sizes from DIABLO components. (**Top**) DIABLO component scores (samples’ projections on the extracted discriminative latent axis) for metabolomics (left) and proteomics (right), stratified by PhenoAge group (*Accelerated* vs *NotAccelerated*). All components show statistically significant differences (Wilcoxon test). The strongest separation is observed in Proteomics Component 1, where *Accelerated* individuals have lower scores. Metabolomics components also show consistent group separation, most notably in Component 2, where *Accelerated* individuals have higher scores. (**Bottom**) Corresponding Cohen’s *d* effect sizes (*Accelerated* vs. *NotAccelerated*) quantify the magnitude and direction of separation between groups. Effect sizes reflect differences in multivariate component scores (latent scores), not raw biomarker levels. Negative values (e.g., Components 1 and 3) indicate higher scores in the *NotAccelerated* group, while positive values (e.g., Component 2 and 4) indicate higher scores in the *Accelerated* group. Overall, proteomics drives stronger separation in Component 1 and 3, whereas metabolites contribute more strongly to Component 2 and 4.

### Molecular features underlying DIABLO components

To identify features contributing to component-level separation, we examined loadings (contribution to component definition) and selection stability (robustness) across cross-validation folds (**Figures 2 and 3**; Supplementary **Tables S1-S4**). **Component 1** was dominated by immune-related proteins (e.g. OSM, TNFRSF1A, IL6), and growth/repair factors (e.g. HGF, TGFA, PLAUR, IGFBP4), together with the inflammatory metabolite glycoprotein acetyls and the glucose:lactate ratio. Functional enrichment highlighted immune activation, inflammatory response, and cytokine signaling pathways. This component represents a pro-inflammatory immune-metabolic axis strongly associated with accelerated aging. **Component 2** was driven by polyunsaturated fatty acids (PUFAs) and cholesteryl esters in medium LDL, alongside proteins involved in receptor tyrosine kinase signalling (ERBB2/3, EGFR, FGFR2), lipid homeostasis (LDLR, TTR, LCAT), complement and coagulation cascades (PROC, SERPINA4, TFPI, MASP1, F7), and appetite/metabolic regulation (AGRP, NPY). Enrichment analysis highlighted ‘complement and coagulation cascades’, and ‘EGFR tyrosine kinase inhibitor resistance’. This component suggests a lipid-vascular regulatory axis. **Component 3** was anchored by glutamine, with additional contribution from the glucose:lactate ratio. Proteomic features included FGF23 (phosphate metabolism), TFRC (iron uptake), and EPO (erythropoiesis), along with APCS, CD160, VCAM1/JAM2 (immune/vascular adhesion), and CRH (neuroendocrine stress signalling). Enrichment analysis highlighted ‘response to nutrient’. This component may represent regulatory nutrient-metabolic processes. **Component 4** was characterized by apolipoproteins (APOM, APOA1, APOC1), APOD and PLTP, together with HDL-related metabolites. Enrichment analysis pointed to cholesterol transport and lipoprotein metabolic processes, suggesting a potentially protective, lipid-regulatory signature enriched in delayed aging. However, both stability scores and effect sizes were small, implying limited robustness. Specific proteins such as top scoring CD300LG may still be of interest, consistent with recent evidence linking this marker to exercise and insulin homeostasis^28^.

**Figure 2.**
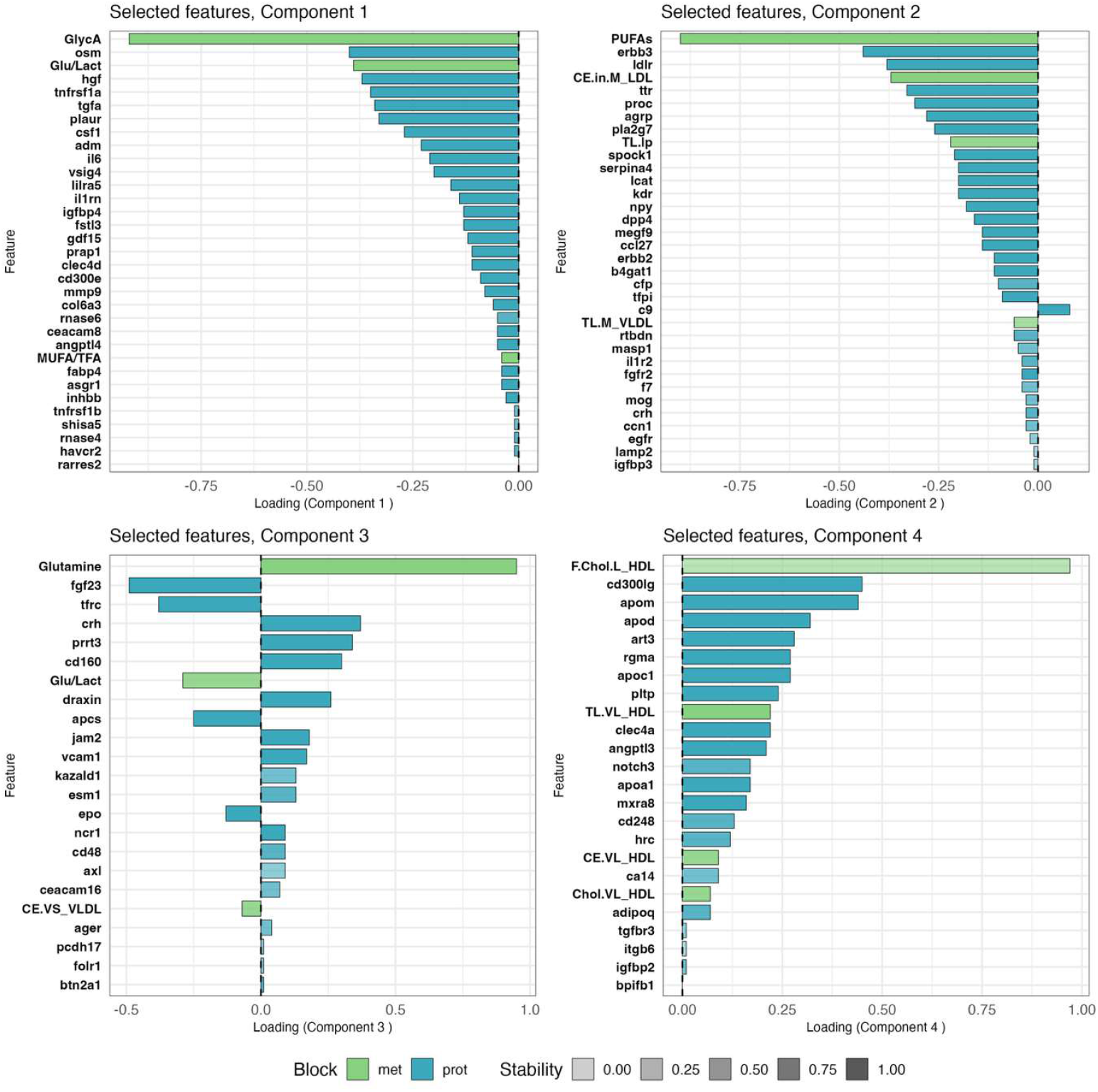
Feature loadings selected by DIABLO across latent components. The loadings of selected proteomic (blue) and metabolomic (green) features for DIABLO Components 1-4 quantify each feature’s contribution to defining the latent component and separating *Accelerated* vs. *NotAccelerated* aging groups. The sign (positive/negative) indicates the direction of association. Transparency of the bars represents stability i.e. selection frequency across cross validation folds. Abbreviated metabolite names are used for readability (e.g., GlycA = Glycoprotein acetyls, TL.M_VLDL = Total lipids in medium VLDL). A complete mapping of abbreviations to full metabolite names is provided in **Table 3**.

**Figure 3.**
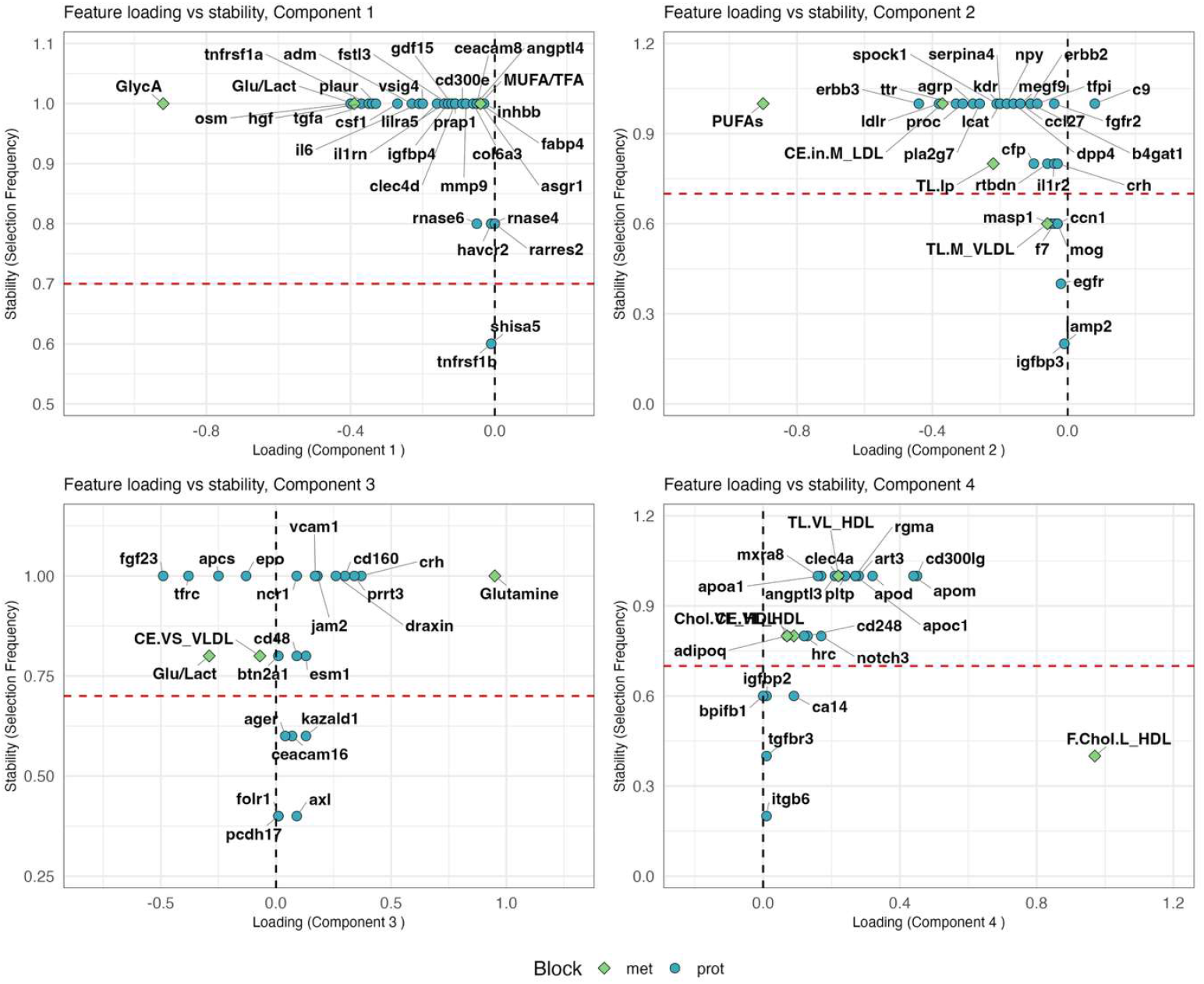
Feature loadings versus stability across DIABLO latent components. Scatterplots show the relationship between feature loadings and selection stability for proteomic (blue) and metabolomic (green) features across DIABLO Components 1-4. The x-axis indicates loading strength and direction, while the y-axis represents selection frequency (stability) across cross validation folds. The red dashed line marks the stability threshold (0.7). Features above this line were selected consistently, highlighting robust contributors such as GlycA and Glu/Lact (Component 1), PUFAs (Component 2), Glutamine and FGF23 (Component 3). Features with both high absolute loadings and high stability are the most biologically interesting, as they combine strong discriminative power with reproducibility. Metabolite abbreviations are defined in **Table 3**.

### Cross-omics relationships and network structure

To visualize integration across omics layers, we plotted correlation circles displaying features from both blocks simultaneously, with stronger components’ contributions from features farther from the origin (Supplementary **Figure S1**). Features clustering together within or across blocks may reflect shared biology. Circos plots (Supplementary **Figure S2a and S2b**) and the relevance network (**Figure 4**) further illustrate interconnections between proteins and metabolites, highlighting potential mechanistic links.

**Figure 4.**
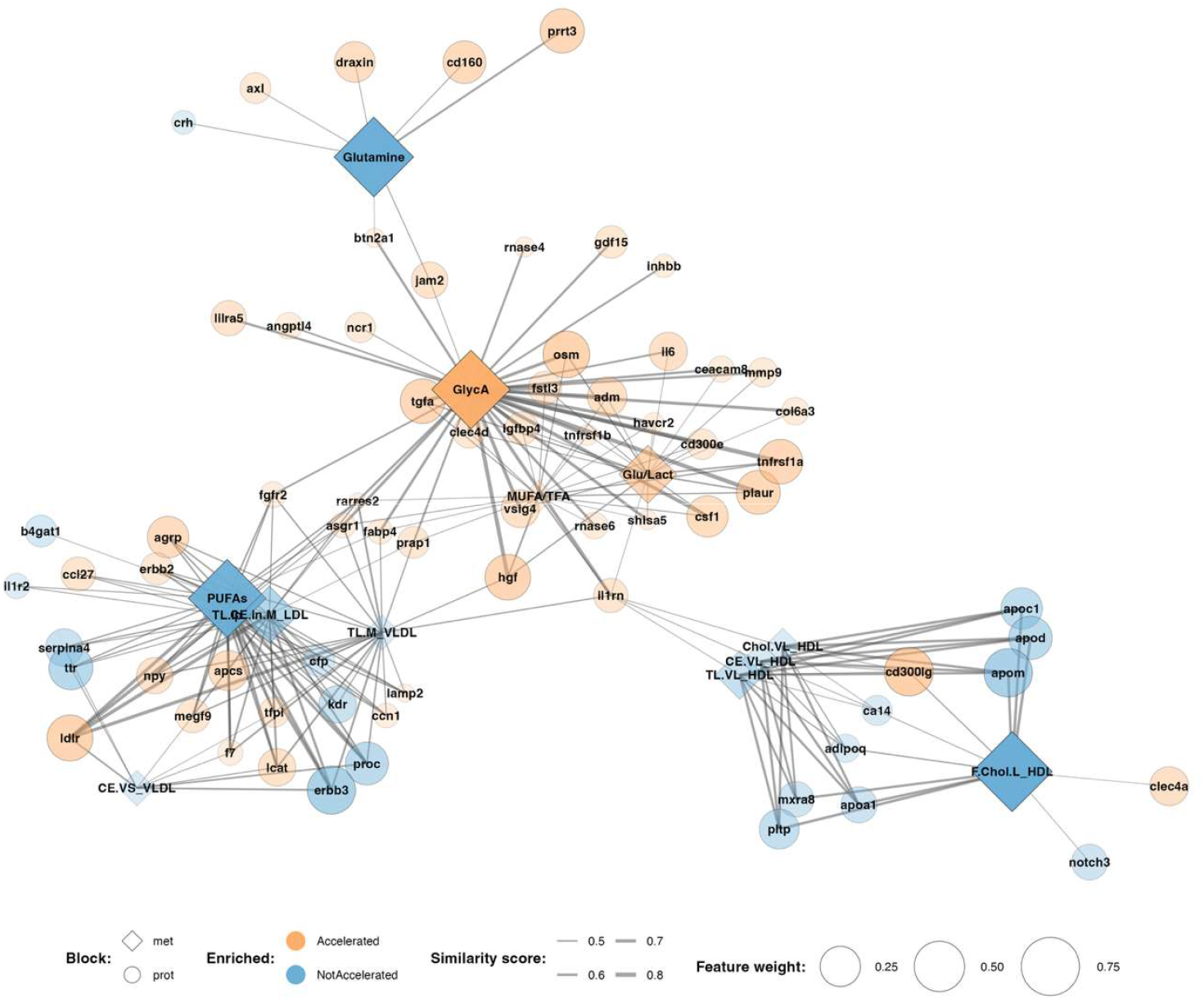
Relevance network of multi-omics features identified by DIABLO across Components 1-4. The network was constructed using a cutoff of |correlation| ≥ 0.4 between selected features. Nodes represent proteomic (circles) and metabolomic (diamonds) variables, with node size scaled to feature weight (importance in the DIABLO model). Node colour denotes relative enrichment in the *Accelerated* (orange) and *NotAccelerated* (blue) PhenoAge groups, based on mean group differences. Edge thickness corresponds to the similarity score (strength of pairwise correlation). Prominent hubs, such as GlycA, Glutamine, PUFAs, and F.Chol.L_HDL, connect multiple features and highlight integrative molecular signatures associated with biological age acceleration. Clustering of nodes reflects correlation structure across omics layers, with features from distinct components predominantly grouping together. This suggests that while component assignment reflects latent factors optimized for group discrimination, the network may represent biological co-regulation across features and blocks (proteomics and metabolomics). Metabolite abbreviations are defined in **Table 3**.

Relevance network analysis revealed a modular structure predominantly aligned with DIABLO components (**Figure 4**). A prominent immune-inflammatory module linked cytokines and growth/repair proteins (e.g. IL6, TNFRSF1A, HGF, IGFBP4, PLAUR, OSM) with glycoprotein acetyls and glucose:lactate ratio. A second module connected lipid metabolites (PUFAs, cholesteryl esters) with receptor tyrosine kinase signaling (ERBB2/3, FGFR2), lipid transport (LDLR, TTR, LCAT), and complement/coagulation proteins (PROC, SERPINA4, TFPI, F7). Smaller subnetworks centred on glutamine and HDL-associated features were also observed. This network represents a correlation structure conditional on the extracted components and emphasizes that DIABLO extracted an interconnected modular structure.

### Relevance Network of multi-omics features associated with Accelerated PhenoAge

### Overlap with established aging and senescence databases

To assess biological relevance, DIABLO-selected proteins were cross-referenced against curated aging resources including the Ageing Gene Database (GenAge)^29^, the Cell Senescence Gene Database (CellAge)^29^, the LongevityMap^29^, the Genetic and Dietary Restriction Database (GenDR)^30^, and the SASP Atlas^31^. Overall, 49 of 100 selected proteins (49%) had at least one aging-related annotation (**Table 2**), exceeding the expected proportion for the human proteome. Overlap was highest in Components 1 (53%) and 2 (60%), consistent with their stronger effect sizes.

**Table 2.**
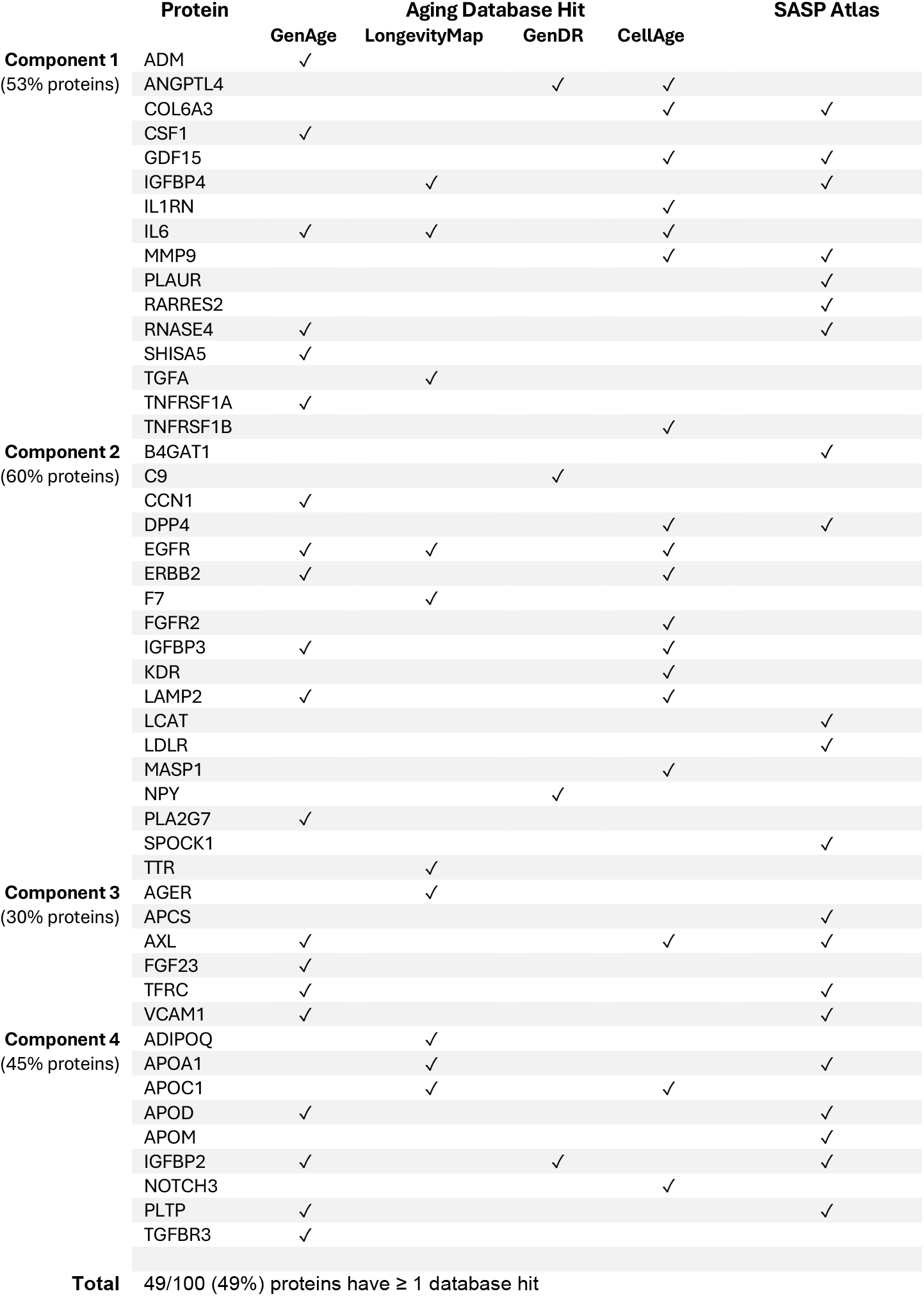
Overlap between DIABLO-selected proteins and established aging databases. Proteins selected within each component were annotated against five major aging resources (GenAge, CellAge, LongevityMap, GenDR, SASP Atlas). Overall, 49 of 100 proteins (49 %) had at least one aging resource hit. Component-specific overlaps were 53 % (Component 1), 60% (Component 2), 30 % (Component 3), and 45 % (Component 4). SASP expression profiles for overlapping proteins are shown in Supplementary **Figure S8**.

**Table 3.**
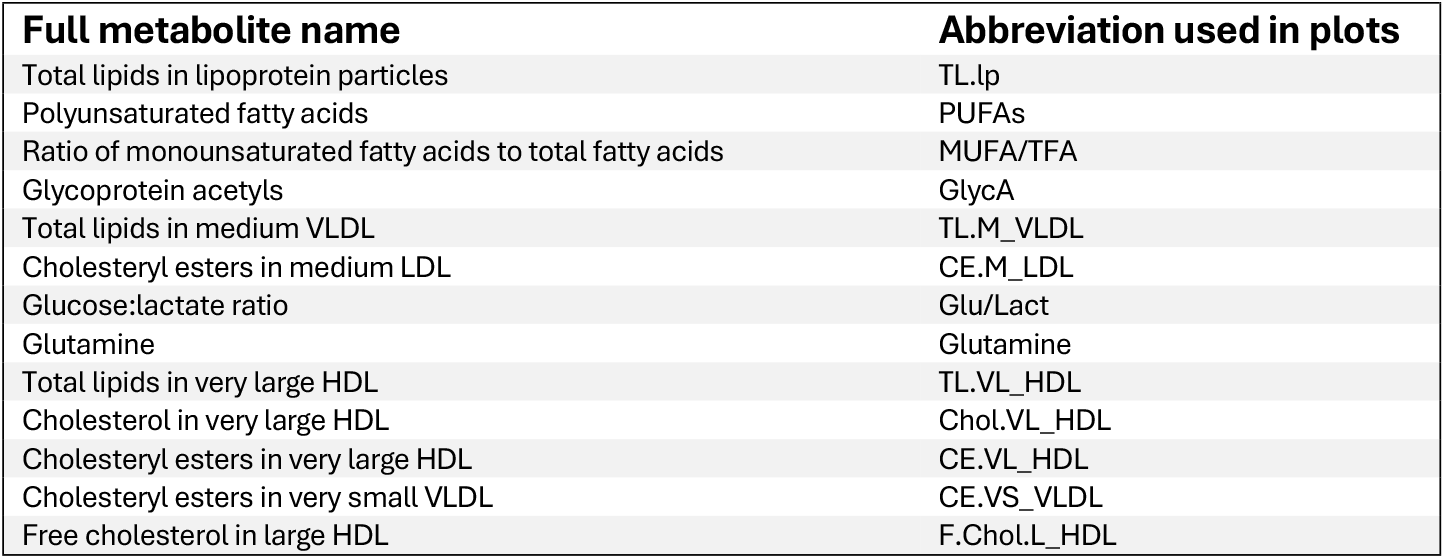
Metabolite abbreviations used in figures. For clarity, long metabolite names were shortened to abbreviated forms in figures (loadings, stability plots, and network representations). Full names correspond to variables measured in the Nightingale NMR metabolomics platform of the UK Biobank.

Notable overlapping proteins included established aging and senescence markers such as GDF15, IL6, MMP9 (Component 1), DPP4, EGFR, ERBB2 (Component 2), AXL, FGF23, and TFRC (Component 3). Among metabolites, several have well-established relevance to aging. Glycoprotein acetyls (GlycA) is a robust marker of systemic inflammation and mortality risk^32^, while polyunsaturated fatty acids (PUFAs) and cholesteryl esters in LDL are central to lipid metabolism and cardiovascular aging^33, 34, 35^. Together, these findings confirm that DIABLO captured features consistent with known hallmarks of aging, providing confidence in the model.

### Novel candidate features

In addition to established aging-related proteins such as GDF15^36^, IL6^37^, MMP9^38^ or FGF23 increasingly recognised as an aging-related biomarker^39, 40^, our analysis identified several proteins with limited links to aging as key contributors to DIABLO components. These proteins include VSIG4, LILRA5, CLEC4D, CD300E, PRAP1 (Component1), MEGF9, SPOCK1, B4GAT1 (Component 2), and PRRT3, DRAXIN, and KAZALD1 (Component 3). PRRT3, with very limited functional annotation, emerged as a top discriminative feature, highlighting a potential novel candidate for future replication and functional investigation. On the metabolite side, features like glucose:lactate ratio (energy metabolism), total lipids in lipoprotein particles, and glutamine with its important role in constructing Component 3 and growing interest in longevity research^41, 42, 43^, highlight metabolic axes for further study in longitudinal and experimental settings.

## Discussion

We integrated plasma proteomics and metabolomics to characterize molecular signatures associated with accelerated biological aging, as quantified by *PhenoAgeAccel*, in a large UK Biobank subcohort. Consistent with prior work^16, 17, 44^, proteomics alone provided strong discrimination between accelerated and slower aging phenotypes. Integrative modeling using DIABLO did not substantially improve classification performance but revealed coordinated multi-omics signatures that enhanced biological interpretability. Overall model performance was comparable to prior omics-based aging studies, which typically report accuracies in the range of 0.65-0.80^17, 45^. The primary advantage of integrative analysis was the identification of biologically interpretable molecular signatures rather than gains in classification accuracy.

Our analysis identified four distinct but interconnected components. The dominant signal was captured by **Component 1**, which reflected immune-inflammatory activation with metabolic dysregulation upregulated in the accelerated aging group. This component aligns with the concept of inflammaging, a key aging process^46, 47, 48^, while also highlighting the importance of immune-metabolic interplay^49^. Several high-ranking features confirmed established links to aging. Among proteins, GDF15^36^, IL6^37^ and MMP9^38^ are considered aging biomarkers associated with inflammation and senescence, while metabolite glycoprotein acetyls (GlycA) robustly predict all-cause mortality^32, 50^. **Component 2** integrated lipid metabolism with vascular and growth factor signalling including PUFAs, cholesteryl esters, receptor tyrosine kinase signalling proteins (ERBB2/3, FGFR2), along with transthyretin (TTR). Beyond its role in thyroid hormone and retinol transport, TTR is implicated in senile systemic amyloidosis (SSA)^51^, increasingly recognized as a contributor to late-life cardiac dysfunction^52, 53^ and has been hypothesised to contribute substantially to mortality among supercentenarians^54^. Its co-selection with lipid and vascular markers suggests that this component may represent an axis that spans cardiometabolic risk in midlife to late-life amyloidosis. **Component 3** identified a smaller module centred on glutamine and several proteins involved in mineral/nutrient regulation (e.g., phosphate via FGF23, iron via TFRC, erythropoiesis via EPO). Glutamine is a major fuel source for rapidly proliferating and immune cells and feeds into the tricarboxylic acid (TCA) cycle via conversion to α-ketoglutarate (AKG)^55^. Recent retrospective human study reported that supplementation with alpha-ketoglutarate based formulation was associated with an average 8-year reduction in biological age estimates after ∼7 months of use^56^. Our observation that glutamine anchors a discriminative aging component suggests that glutamine-AKG metabolism may represent a broader nutrient-energy axis relevant to aging. **Component 4** grouped features predominantly enriched in not accelerated group including HDL-associated lipids and protective apolipoproteins. CD300LG, strongly contributing to this component, was recently described as an exercise-responsive exerkine linked to glucose homeostasis^28^, suggesting that this axis may reflect favourable metabolic adaptations rather than core aging processes. Even though modest effect sizes are typical of gerontology research^57^, due to very small effect sizes this component should be interpreted with caution.

Complementary visualizations supported the component-level interpretations. Correlation circles showed clustering of proteomic and metabolomic features across blocks (Supplementary **Figure S1**), while circos plots highlighted cross-omics associations (Supplementary **Figures S2a and S2b**). The relevance network revealed modular structure, with hubs linking proteins and metabolites within shared pathways (**Figure 4**). Together, they reinforce the biological plausibility of the selected features and show how integrative methods improve interpretability beyond feature loadings alone. Importantly, nearly half of DIABLO-selected proteins overlapped curated aging and senescence databases, with enrichment strongest in the components showing the largest effect sizes. This supports the biological validity of the identified signatures while also highlighting proteins and pathways not yet well-characterized in the context of aging. Several novel or poorly annotated proteins, such as PRRT3 and DRAXIN (Component 3), warrant further investigation. Methodologically, our results illustrate that the value of multi-omics integration lies less in gains in predictive performance and more in the ability to uncover structured, biologically coherent multi-omics signatures. Although metabolites showed weaker standalone performance, they played a critical anchoring role within integrated components, linking protein-level changes to underlying metabolic states and enhancing mechanistic interpretability.

This study has several limitations including cross-sectional design preventing causal inference, and reliance on plasma measurements, which may not capture tissue-specific aging processes. Although novel candidates were highlighted, independent and statistically rigorous validation and functional follow-up is essential to establish their relevance. We have included 248 metabolites (NMR) and 2,920 proteins (Olink) implicated in age-related disease, but untargeted metabolomics, broader proteomics, or other platforms may reveal additional signatures. Additionally, more extensive adjustments for covariates such as smoking, alcohol use, physical activity, obesity, medication, socio-economic status, or stratified analyses, should be considered. Additional omics layers, such as transcriptomics or epigenetics, may further refine the results.

Because PhenoAge is derived from clinical measures including inflammatory and metabolic biomarkers, we excluded metabolites directly used in its calculation (i.e., creatinine, albumin, and glucose) from the omics blocks prior to integration analyses to minimise circularity (see Methods). Nevertheless, it is important to recognise that multi-omics data capture biologically correlated features within the same domains. In this context, the identification of immune-metabolic molecular signatures is biologically consistent but also partly expected. Our findings suggest that *PhenoAgeAccel* is associated with immune-metabolic dysregulation extending beyond the specific variables used to construct the phenotypic aging measure. Future work evaluating multi-omics signatures against aging metrics that do not explicitly include inflammatory biomarkers (e.g., epigenetic clocks) would further clarify the extent to which these patterns reflect biological convergence versus outcome-specific structure.

## Conclusions

In summary, our integrative analysis identified distinct immune-metabolic, lipid-vascular, nutrient-metabolic, and HDL-associated signatures linked to PhenoAge acceleration. While proteomics captures the dominant discriminative signal, metabolomics provides essential contextual information so that we see not just which proteins change, but why those changes may reflect underlying metabolic, nutrient, and energy pathways. These findings reinforce the multifactorial nature of aging and highlight coordinated molecular pathways that may inform future mechanistic studies and interventions aimed at extending healthspan.

## Methods (Summary)

### Study population

This study utilised the UK Biobank^25^, a large prospective population-based cohort of ∼500,000 participants. We focused on a randomly selected subset with both proteomic and metabolomic measurements available from blood samples collected at baseline. Within this subset, individuals with complete baseline data for the nine clinical blood biomarkers required to calculate PhenoAge^5^ were included (n = 22,711). Samples with >20% of missing values in proteomic assays were excluded, resulting in a final sample of 18,632 participants. This research has been conducted using the UK Biobank Resource under Application Number 438487.

### Outcome definition

Biological age was estimated using PhenoAge^5^, a clinically grounded biomarker-based measure derived from nine routine blood biomarkers and chronological age. PhenoAge acceleration (*PhenoAgeAccel*) was calculated as the residual from regressing PhenoAge on chronological age, reflecting faster or slower aging than expected. For multivariate analyses, individuals in the upper and lower 20% of the *PhenoAgeAccel* distribution were selected to enhance biological contrast between accelerated and slower aging phenotypes.

### Omics data

Plasma proteomics was measured using Olink proximity extension assays, comprising 2,920 proteins after quality control. Metabolomic profiling was performed using the Nightingale Health targeted NMR platform, yielding 248 metabolites after exclusion of biomarkers used in PhenoAge calculation. Missing values were imputed using k-nearest neighbors, and metabolite data were log1p-transformed. Chronological age and sex were regressed out of omics data prior to multivariate modeling.

### Single-omics analysis

Sparse partial least squares discriminant analysis (sPLS-DA) was applied separately to proteomic and metabolomic data to assess their individual ability to discriminate accelerated from slower aging. Model performance was evaluated using cross-validation and balanced error rates.

### Multi-omics integration

Proteomic and metabolomic data were jointly modeled using DIABLO, a supervised multiomics integration framework within the mixOmics package. DIABLO identifies correlated features across data modalities while optimizing discrimination of the outcome^27^. The number of components and features retained per component were selected using repeated cross-validation. Model performance was assessed using balanced error rates and area under the ROC curve in both training and independent test sets.

### Interpretation and validation

Biological relevance was evaluated using component score effect sizes, feature stability across cross-validation folds, functional enrichment analyses, and cross-referencing of selected proteins with curated aging and senescence databases. Network and correlation-based visualizations were used to explore cross-omics relationships.

**Detailed methodological descriptions, including model specification, parameter tuning, and supplementary analyses, are provided in the Supplementary Material**.

## Data Availability

All data produced are available online at UK Biobank.

## Acknowledgments

We thank the members of the Genomics of Ageing and Rejuvenation Lab, Department of Inflammation and Ageing, College of Medicine and Health, University of Birmingham, for helpful discussions. This research received no specific grant from any funding agency in the public, commercial, or not-for-profit sectors.

## Conflict of interest

JPM is CSO of YouthBio Therapeutics, an advisor/consultant for the BOLD Longevity Growth Fund and NOVOS, and the founder of Magellan Science Ltd, a company providing consulting services in longevity science.

## Notes

### Funding Statement

This study did not receive any funding.

### Author Declarations

This research has been conducted using the UK Biobank Resource under Application Number 438487.

