## Supplementary material for "Integrative Proteomic and Metabolomic Signatures of Accelerated PhenoAge in the UK Biobank"

### Methods

#### 1. Study population

This study utilised the UK Biobank[1], a large prospective population-based cohort of ~500,000 participants. We focused on a randomly selected subset with both proteomic and metabolomic measurements available from blood samples collected at baseline. Within this subset, individuals with complete baseline data for the nine clinical blood biomarkers required to calculate PhenoAge were included (n = 22,711). Samples with >20% of missing values in proteomic assays were excluded, resulting in a final sample of 18,632 participants. Data were accessed under UK Biobank Application Number 438487 and all participants provided informed consent.

#### 2. Data Sources and pre-processing

##### 2.1.Proteomics

Plasma proteomic profiles were generated using Olink's Proximity Extension Assay (PEA) technology, covering 2,923 proteins. Abundances were reported as Normalised Protein eXpression (NPX) values on a log2 scale, representing relative concentrations. The NPX values were normalised and quality-filtered prior to the release[2, 3]. From the available proteins, those with >25% of missing values (n = 3) were excluded, leaving 2,920 proteins for analysis. Remaining missing values were imputed using the K-Nearest Neighbors (KNN) algorithm.

##### 2.2.Metabolomics

Metabolomic data were obtained from the targeted Nightingale Health nuclear magnetic resonance (NMR) platform, measuring 251 absolute concentrations from EDTA plasma. Quality control and normalization were performed prior to release[4]. Metabolites directly used to calculate PhenoAge (creatinine, albumin, and glucose) were excluded from the final model, leaving 248 metabolites. Missing values were imputed using KNN algorithm, and metabolite values were log1p-transformed to approximate a normal distribution.

The large sample size, together with platform specific QC, including batch effect assessments and normalization, reduces the impact of technical variation and increases the robustness of downstream analyses.

### 3. Outcome

PhenoAge was calculated as described by Levine et al. (2018)[5], combining nine clinical blood biomarkers (albumin, creatinine, glucose, log-transformed C-reactive protein, lymphocyte %, mean cell volume, red cell distribution width, alkaline phosphatase, and white blood cell count) with chronological age.

Step 1. Estimating 10-year mortality risk

For each individual $j$, the 10-year (120-month) mortality risk was estimated from the cumulative distribution function (CDF) of a parametric Gompertz proportional hazards model:

$$MortalityScore_j = CDF(120, x_j) = 1 - \exp\left(\frac{-\exp(x_j \cdot \beta) \cdot (\exp(120 \cdot \gamma) - 1)}{\gamma}\right) \quad (1)$$

where: $x_j \cdot \beta$ is the linear combination of biomarker values and their regression coefficients (**Supplementary Table S5)** derived from a penalized Cox model of aging-related mortality. The shape parameter was set at $\gamma = 0.0076927$, as estimated in the original study[5].

Step 2. Mapping risk to biological age

The estimated mortality risk was then mapped back to an equivalent chronological age, representing the phenotypic biological age, using the inverse of the Gompertz function fitted on age alone:

$$PhenoAge_j = 141.50225 + \frac{\ln(-0.00553 * \ln(1 - MortalityScore_j))}{0.090165} \quad (2)$$

This value corresponds to the age at which an individual with reference biomarker values would have the same predicted 10-year mortality risk as individual $j$.

PhenoAge acceleration (*PhenoAgeAccel*) was calculated as the residual from regressing PhenoAge on chronological age. It reflects the extent to which an individual's biological age (PhenoAge) is higher or lower than expected for their chronological age within our study cohort.

To facilitate classification task, a binary outcome variable was defined by categorizing individuals as *Accelerated* (*PhenoAgeAccel* > 0) and *NotAccelerated* (*PhenoAgeAccel* <= 0).

### 4. Single-omics analysis

To characterize proteomic and metabolomic datasets and assess their individual contributions to accelerated aging, we first examined each omics layer separately.

#### 4.1. Univariate analysis

Differential analysis of plasma proteins and metabolites was performed to identify features associated with *PhenoAgeAccel*, comparing the *Accelerated* vs *NotAccelerated* groups (**Supplementary Table S6-S7**). Linear models were fitted using the *limma* R package, with age and sex as covariates. False discovery rate (FDR) was controlled using the Benjamini-Hochberg correction, and features with adjusted p-value < 0.05 were considered significant. In addition, absolute log2 fold-change (FC) thresholds of 0.2 (proteins) and 0.04 (metabolites) were applied, reflecting the lower dynamic range of metabolomic data. Significant features were subjected to functional enrichment analysis (Gene Ontology and KEGG) with a custom background, as described in the *Feature importance analyses* (section 5.4.).

#### 4.2. Unsupervised multivariate analysis

Principal Component Analysis (PCA) was used to explore variance structure within each omics dataset before integration. To enhance group separation, analyses were restricted to individuals with extreme *PhenoAgeAccel* values (≤ $20^{th}$ percentile or ≥ $80^{th}$ percentile). This subset of 7,579 samples showed clearer separation of *Accelerated* vs *NotAccelerated* groups compared to the full dataset, likely due to increased phenotypic contrast and reduced overlap between borderline cases.

#### 4.3. Discriminant analysis

We next assessed the predictive capacity of each omics block using sparse Partial Least Squares Discriminant Analysis (sPLS-DA, *mixOmics* R package). Analyses were performed on the same subset of 7,579 individuals with extreme *PhenoAgeAccel* values. Chronological age and sex were regressed out prior to model fitting to reduce confounding. The optimal number of components was chosen based on classification error rate estimated by 5-fold cross-validation. Grid search (*tune()* function, 5-fold CV, 'max.dist' prediction distance) was used to optimize the number of variables retained per component. The final proteomics model selected (30, 90, 80, 90, 40) proteins across five components (**Supplementary Figure S3**). In contrast, the metabolomics model required eight components, retaining (6, 10, 9, 25, 15, 10, 6, 35) features, respectively (**Supplementary Figure S4**).

### 5. Multi-omics integration

#### 5.1. Analytical method selection and model framework

Multi-omics approaches can capture more subtle signals and cross-omics associations than single-omics analyses. However, integration is challenging due to high dimensionality, risk of overfitting, multicollinearity that can generate spurious associations, and biological as well as technical heterogeneity. Selecting an analytical method that could accommodate these issues while addressing the study aim was therefore critical.

Our objective was to identify a multi-omics signature, correlated features across proteomics and metabolomics, that also discriminated the outcome (*Accelerated* vs *NotAccelerated* aging). We selected Data Integration Analysis for Biomarker Discovery using Latent Variable Approaches for Omics Studies (DIABLO)[6] from the *mixOmics* R package. DIABLO is generally recommended when outcome information is available and has outperformed other integration methods in tasks such as cancer type classification[7]. Its latent variable framework, combined with feature selection, extracts biologically meaningful, compact signatures that are robust and interpretable[8]. The method uses matrix factorization for dimensionality reduction, ridge regularization to stabilize estimation in high-dimensional settings, and lasso regularization for variable selection. The classification-oriented framework enables cross-validation and evaluation on independent test data, assessing model generalizability.

We applied DIABLO to 7,579 individuals with extreme *PhenoAgeAccel* values ($\leq$ Q20 or $\geq$ Q80). This subset was split into stratified Train (70%) and Test (30%) sets. Covariates age and sex were regressed out from omics data prior to model fitting.

#### 5.2.Tuning and parameters choice

The DIABLO design matrix controls the balance between maximizing correlations across the datasets and discrimination of the outcome. Because our primary aim was to identify features predictive of accelerated aging, we used a full-weighted design with a weight = 1 between each block and the outcome, and a lower weight = 0.1 for proteomics and metabolomics connection.

To choose the number of components, we used repeated 5-fold cross-validation (5 repeats) and evaluated classification error via *perf()* function (**Supplementary Figure S5**). Based on these results, we selected four components, the 'max.dist' prediction distance, and 'WeightedVote' as the prediction scheme.

To optimize the number of retained variables per block and per component, we performed a grid search using *tune.block.splsda()* function with 5-fold cross-validation. The final model retained a minimal set of features while preserving classification performance, yielding (30, 30, 20, 20) proteins and (3, 4, 3, 4) metabolites across four components.

#### 5.3.Model validation

Internal validation was performed using *perf()* function with 5-fold cross-validation, 'max.dist' prediction distance, and 'WeightedVote' scheme. Overall and balanced error rates were similar, indicating that class imbalance was not a concern. ROC curves and AUC values were inspected to confirm discriminative ability (**Supplementary Figure S6**). Model generalizability was assessed on the independent Test dataset. Visualization was integral to model assessment. Component score plots illustrated class separation (**Supplementary Figure S7**). Loading plots (**Main text Figure 2**) and correlation circles (**Supplementary Figure S1**) enabled inspection of variable contributions and inter-relationships. We also compared DIABLO to single-omics sPLS-DA models, assessing not only classification performance but also interpretability and biological plausibility, including functional enrichment of selected features.

#### 5.4.Feature importance analysis

Selected DIABLO features were subjected to enrichment analysis. For proteins, Gene Ontology (GO) and Kyoto Encyclopedia of Genes and Genomes (KEGG) enrichment was performed using *clusterProfiler* R package, with the background defined as all QC-passed proteins. Adjusted p-values $< 0.05$ (Benjamini-Hochberg) were considered significant.

Graphical outputs including relevance network and correlation circle plots provided insights into relationships between omics features. Features with the highest absolute loadings (strongest contribution to component definition), and highest stability scores (consistent selection across folds) were prioritized as candidate biomarkers (**Main text Figure 3**).

**Software and Reproducibility**

All analyses were performed in R v. 4.3.1. Workflow was managed using R Markdown and *renv* for reproducibility. GitHub repository for code available at: https://github.com/KamilaBloch/UKB_Project

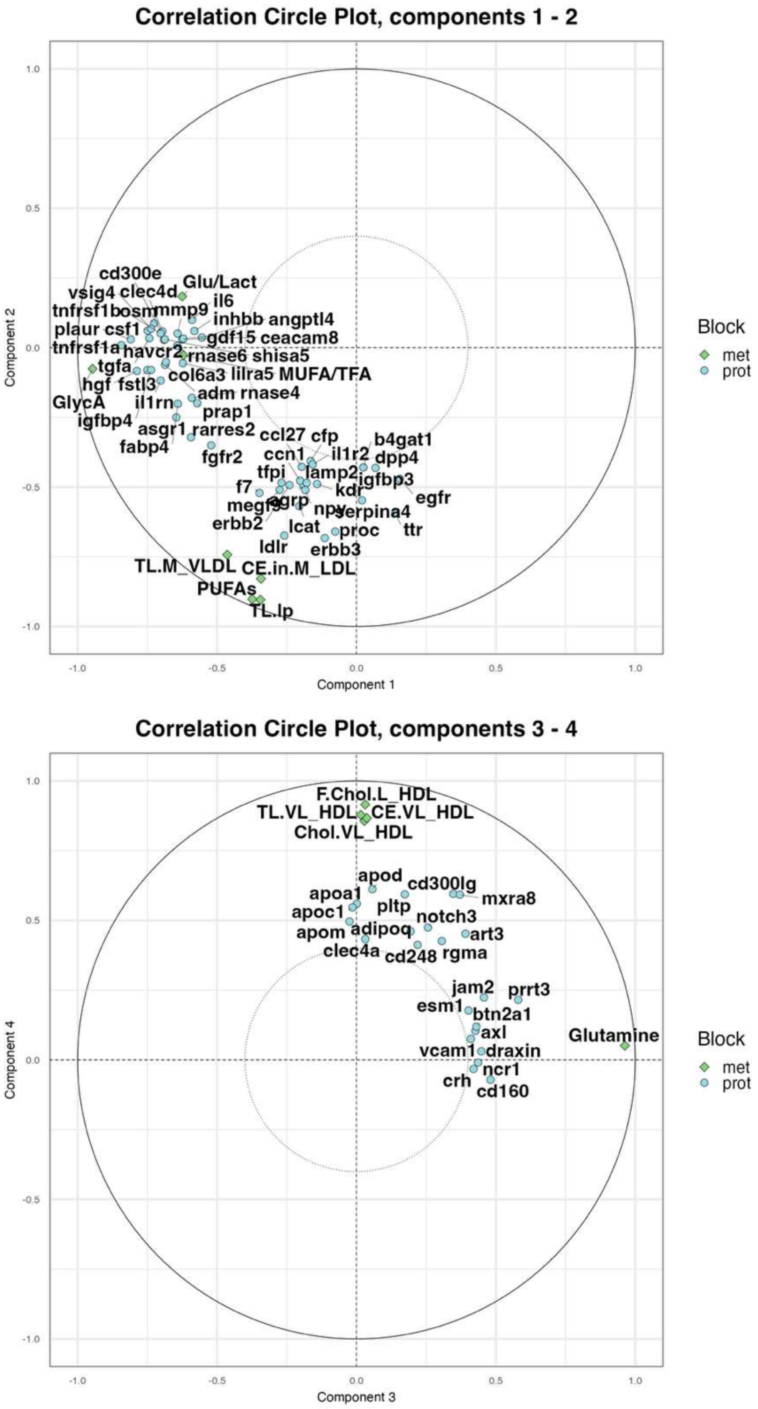

**Figure S1. Correlation circle plots of DIABLO components (block integration).** The plots show the projection of proteomic (blue) and metabolomic (green) features onto DIABLO latent components 1–2 (top) and 3–4 (bottom). Distances from the origin indicate the strength of association between each feature and the components. The outer circle corresponds to a correlation of 1, and the inner dashed circle to 0.4. Features closer to the outer circle are strongly represented in the component space, while those near the origin contribute weakly. Inflammatory and glycoprotein-related features (e.g., GlycA, IL6, TNFRSF1A) dominate Component 1, whereas lipids (e.g. PUFAs, TL.lp, F.Chol.L_HDL, CE.VL_HDL) are strongly represented on Components 2 and 4. Glutamine dominates Component 3. Clusters of features pointing in similar directions indicate groups of likely correlated variables and may reflect shared biological pathways. Features far from the origin and aligned with component axes are the most influential for discrimination. Metabolite abbreviations are defined in **Table S9**.

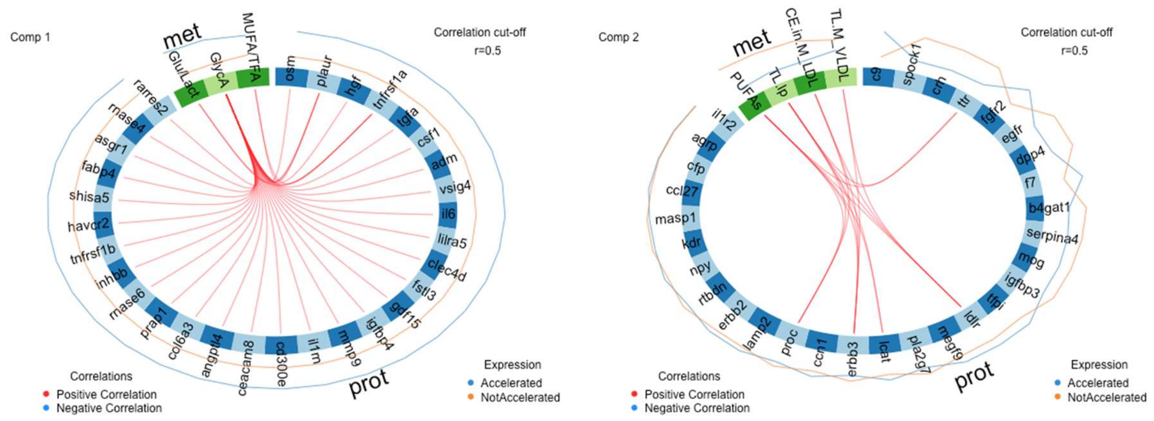

**Figure S2a. Circos plots, cross-block correlations between selected metabolites and proteins for Components 1 and 2.** The circos plots display correlations (**threshold *r* ≥ 0.5**) between metabolite (green) and protein (blue) features selected by DIABLO. Red lines indicate positive correlations. The outer coloured tracks (blue = *Accelerated*, orange = *NotAccelerated*) represent relative group levels. In Component 1, GlycA and MUFA/TFA show strong positive correlations with a network of inflammatory and signalling proteins (e.g., IL6, TNFRSF1A, PLAUR), suggesting a coordinated metabolic–inflammatory axis linked to accelerated aging. In Component 2, lipid measures (PUFAs, CE.M_LDL, TL.M_VLDL) correlate positively with PROC, ERBB3, LCAT, LDLR, and TTR proteins. Metabolite abbreviations are defined in **Table S9**.

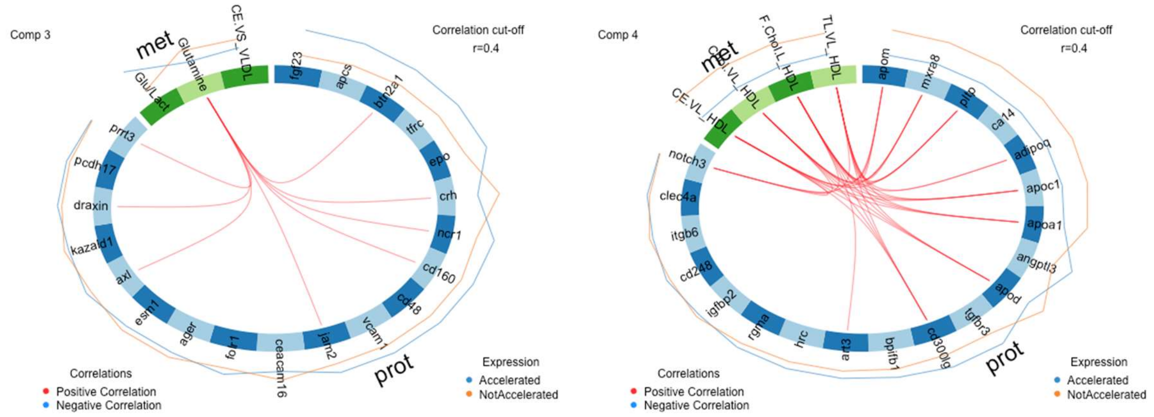

**Figure S2b. Circos plots, cross-block correlations between selected metabolites and proteins for Components 3 and 4.** The circos plots display correlations (**threshold *r* ≥ 0.4**) between metabolite (green) and protein (blue) features selected by DIABLO. Red lines indicate positive correlations. The outer coloured tracks (blue = *Accelerated*, orange = *NotAccelerated*) represent relative group levels. For Component 3, the metabolite glutamine and the glucose:lactate ratio showed positive correlation to PRRT3, DRAXIN, AXL, JAM2, CD160, NCR1, CRH, and BTN2A1. For Component 4, HDL-related metabolites correlated with NOTCH3, ART3, CD300LG, APOD, APOA1, APOC1, ADIPOQ, PLTP, MXRA8, and APOM proteins. Metabolite abbreviations are defined in **Table S9**.

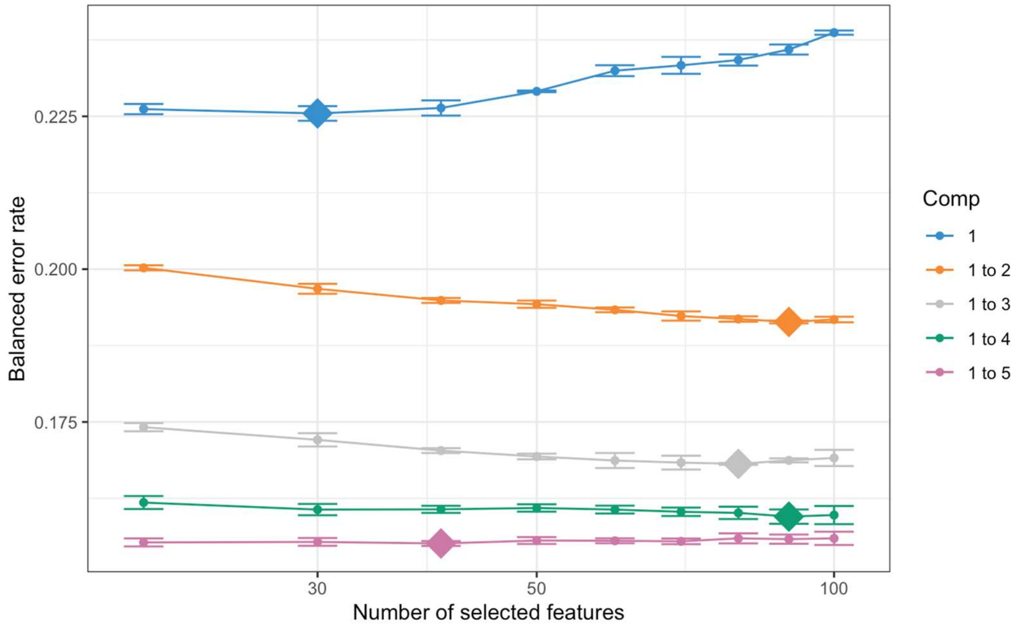

**Figure S3. Tuning of proteomic sPLS-DA model.** Classification error rates (balanced error rate, BER) were estimated by 5-fold cross-validation to determine the optimal number of components and subsequently number of features to retain per component. The final model selected five components with (30, 90, 80, 90, 40) proteins.

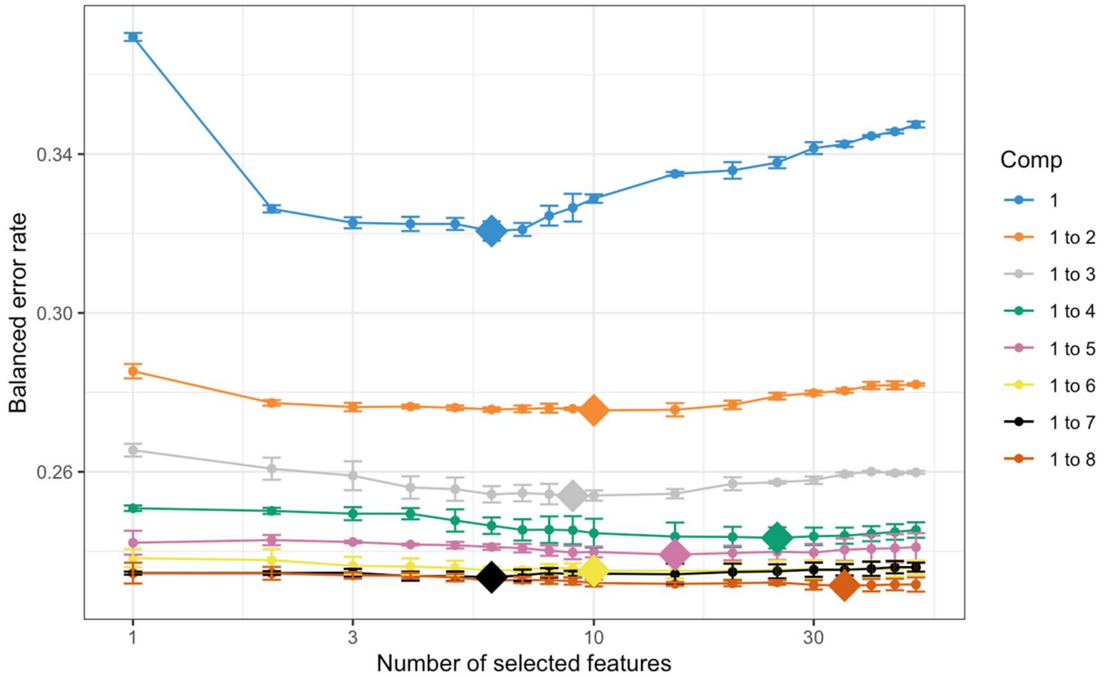

**Figure S4. Tuning of metabolomic sPLS-DA model.** Classification error rates (balanced error rate, BER) were estimated by 5-fold cross-validation to determine the optimal number of components and subsequently number of features to retain per component. The final model selected eight components with (6, 10, 9, 25, 15, 10, 6, 35) metabolites.

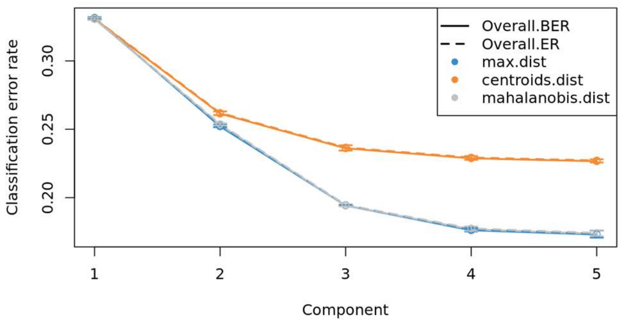

**Figure S5. Tuning the number of components in DIABLO.**
Classification error rates are shown across 1–5 components using different prediction distances (maximum distance, centroid distance, and Mahalanobis distance). Performance improves with additional components, plateauing after four, indicating that a 4-component model provides a good trade-off between complexity and classification accuracy.

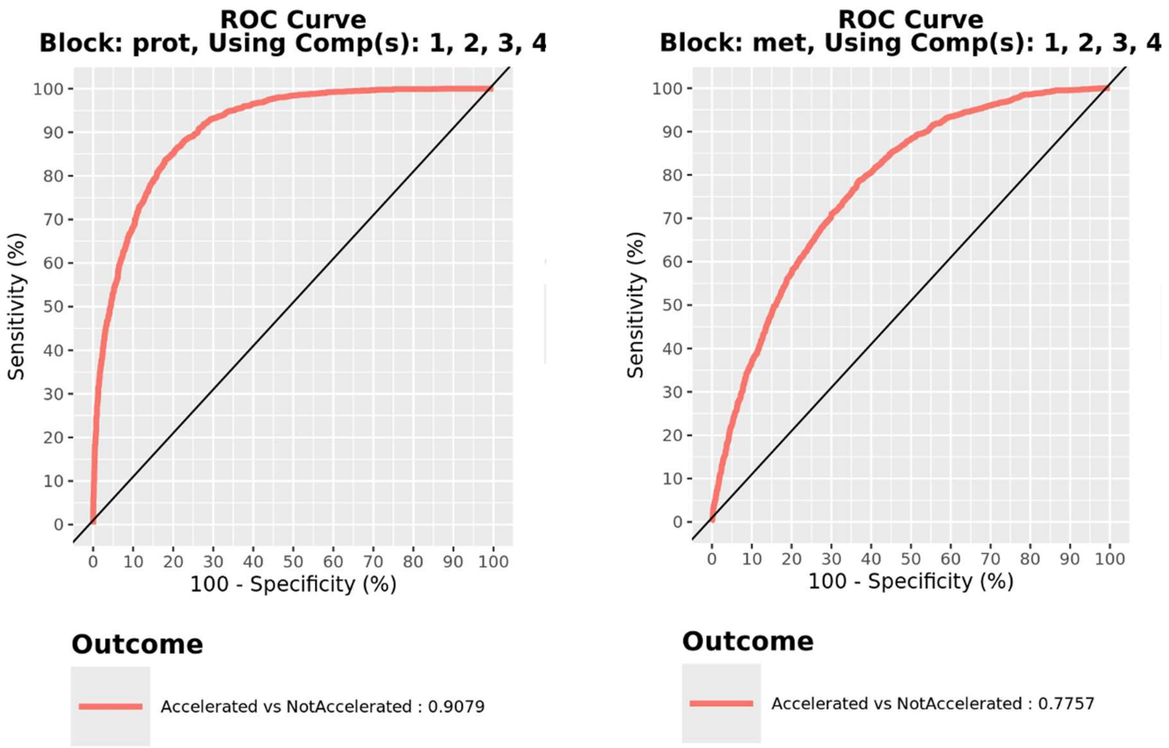

**Figure S6. Receiver Operating Characteristic (ROC) curves for DIABLO components in proteomic (left) and metabolomic (right) blocks.** ROC curves were generated using components 1–4 from the DIABLO model to evaluate the ability of each omics block to discriminate *Accelerated* vs. *NotAccelerated* groups. The proteomic block achieved an area under the curve (AUC) of 0.91, indicating strong classification performance, whereas the metabolomic block achieved a lower AUC of 0.78, consistent with weaker discriminative capacity of metabolites when used alone.

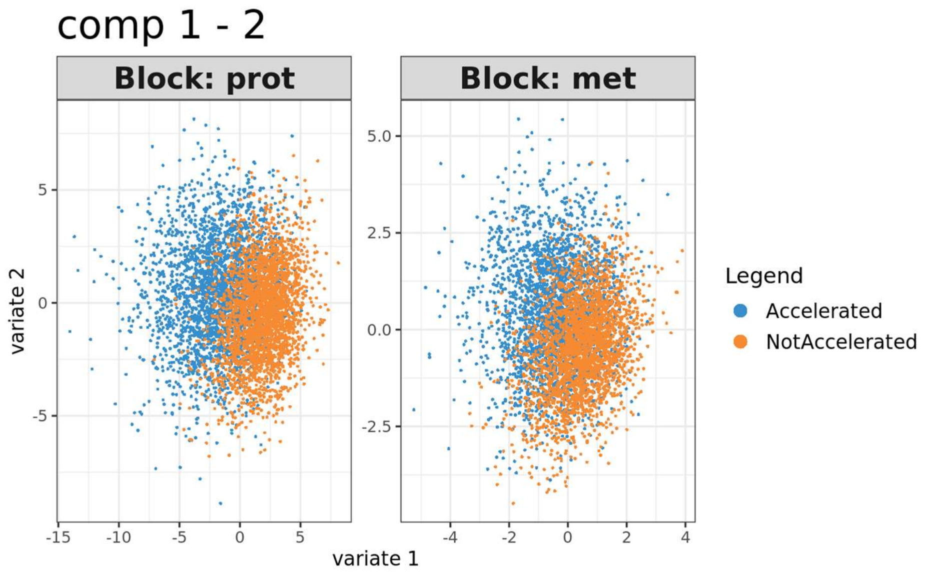

**Figure S7. Sample scatterplots of DIABLO component scores for proteins and metabolites.** Scores from components 1 and 2 are shown for the proteomic (left) and metabolomic (right) blocks. Each point represents one participant, coloured by aging group (*Accelerated* vs. *NotAccelerated*). The plots illustrate partial separation between groups, with proteomic scores showing stronger discrimination than metabolomic scores.

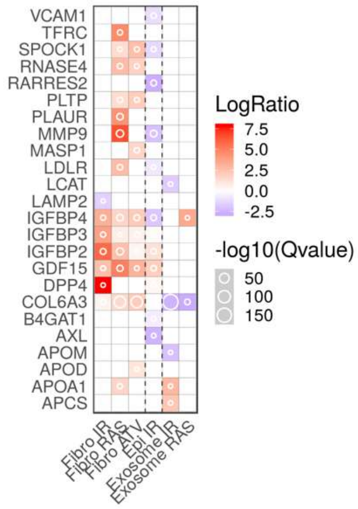

**Figure S8. SASP Atlas profiles of DIABLO-selected proteins.** Proteins identified by DIABLO were cross-referenced with the SASP Atlas to determine whether they are secreted during cellular senescence. The heatmap shows the $\log_2$ fold-change (LogRatio) in protein abundance between senescent and control cells originating from multiple senescence inducers and cell types: fibroblast irradiation (Fibro IR), fibroblast RAS activation (Fibro RAS), fibroblast atazanavir-induced (Fibro ATV), epithelial irradiation (Epi IR), and exosome fractions from irradiated and RAS-induced cells (Exosome IR, Exosome RAS). The colour scale represents the direction and magnitude of change (red = upregulated; blue = downregulated), while circle size denotes $-\log_{10}$(Q-value), reflecting the level of statistical significance reported in the SASP Atlas dataset. Underlying $\log_2$ fold-change data for these profiles are available in **Supplementary Table S8**.

| Selected features, component 1 | Loading | Stability |
|---|---|---|
| Glycoprotein.acetyls | -0.92 | 1 |
| Glucose.lactate | -0.39 | 1 |
| Ratio.of.monounsaturated.fatty.acids.to.total.fatty.acids | -0.04 | 1 |
| osm | -0.4 | 1 |
| hgf | -0.37 | 1 |
| tnfrsf1a | -0.35 | 1 |
| tgfa | -0.34 | 1 |
| plaur | -0.33 | 1 |
| csf1 | -0.27 | 1 |
| adm | -0.23 | 1 |
| il6 | -0.21 | 1 |
| vsig4 | -0.2 | 1 |
| lilra5 | -0.16 | 1 |
| il1rn | -0.14 | 1 |
| fstl3 | -0.13 | 1 |
| igfbp4 | -0.13 | 1 |
| gdf15 | -0.12 | 1 |
| clec4d | -0.11 | 1 |
| prap1 | -0.11 | 1 |
| cd300e | -0.09 | 1 |
| mmp9 | -0.08 | 1 |
| col6a3 | -0.06 | 1 |
| rnase6 | -0.05 | 0.8 |
| angptl4 | -0.05 | 1 |
| ceacam8 | -0.05 | 1 |
| fabp4 | -0.04 | 1 |
| asgr1 | -0.04 | 1 |
| inhbb | -0.03 | 1 |
| tnfrsf1b | -0.01 | 0.6 |
| havcr2 | -0.01 | 0.8 |
| rnase4 | -0.01 | 0.8 |
| shisa5 | -0.01 | 0.6 |
| rarres2 | 0 | 0.8 |

**Table S1**. List of proteomic and metabolomic features selected in DIABLO Component 1. For each feature, the loading indicates its weight in defining the component, while stability reflects the frequency with which the feature was selected across cross-validation folds, providing a measure of robustness.

| Selected features, component 2 | Loading | Stability |
|---|---|---|
| Polyunsaturated.fatty.acids | -0.9 | 1 |
| Cholesteryl.esters.in.medium.LDL | -0.37 | 1 |
| Total.lipids.in.lipoprotein.particles | -0.22 | 0.8 |
| Total.lipids.in.medium.VLDL | -0.06 | 0.6 |
| erbb3 | -0.44 | 1 |
| ldlr | -0.38 | 1 |
| ttr | -0.33 | 1 |
| proc | -0.31 | 1 |
| agrp | -0.28 | 1 |
| pla2g7 | -0.26 | 1 |
| spock1 | -0.21 | 1 |
| serpina4 | -0.2 | 1 |
| kdr | -0.2 | 1 |
| lcat | -0.2 | 1 |
| npy | -0.18 | 1 |
| dpp4 | -0.16 | 1 |
| ccl27 | -0.14 | 1 |
| megf9 | -0.14 | 1 |
| erbb2 | -0.11 | 1 |
| b4gat1 | -0.11 | 1 |
| cfp | -0.1 | 0.8 |
| tfpi | -0.09 | 1 |
| c9 | 0.08 | 1 |
| rtbdn | -0.06 | 0.8 |
| masp1 | -0.05 | 0.6 |
| fgfr2 | -0.04 | 1 |
| il1r2 | -0.04 | 0.8 |
| f7 | -0.04 | 0.6 |
| mog | -0.03 | 0.6 |
| ccn1 | -0.03 | 0.6 |
| crh | -0.03 | 0.8 |
| egfr | -0.02 | 0.4 |
| igfbp3 | -0.01 | 0.2 |
| lamp2 | -0.01 | 0.2 |

**Table S2**. List of proteomic and metabolomic features selected in DIABLO Component 2. For each feature, the loading indicates its weight in defining the component, while stability reflects the frequency with which the feature was selected across cross-validation folds, providing a measure of robustness.

| Selected features, component 3 | Loading | Stability |
|---|---|---|
| Glutamine | 0.95 | 1 |
| Glucose.lactate | -0.29 | 0.8 |
| Cholesteryl.esters.in.very.small.VLDL | -0.07 | 0.8 |
| fgf23 | -0.49 | 1 |
| tfrc | -0.38 | 1 |
| crh | 0.37 | 1 |
| prrt3 | 0.34 | 1 |
| cd160 | 0.3 | 1 |
| draxin | 0.26 | 1 |
| apcs | -0.25 | 1 |
| jam2 | 0.18 | 1 |
| vcam1 | 0.17 | 1 |
| esm1 | 0.13 | 0.8 |
| epo | -0.13 | 1 |
| kazald1 | 0.13 | 0.6 |
| axl | 0.09 | 0.4 |
| cd48 | 0.09 | 0.8 |
| ncr1 | 0.09 | 1 |
| ceacam16 | 0.07 | 0.6 |
| ager | 0.04 | 0.6 |
| btn2a1 | 0.01 | 0.8 |
| folr1 | 0.01 | 0.4 |
| pcdh17 | 0.01 | 0.4 |

**Table S3**. List of proteomic and metabolomic features selected in DIABLO Component 3. For each feature, the loading indicates its weight in defining the component, while stability reflects the frequency with which the feature was selected across cross-validation folds, providing a measure of robustness.

| Selected features, component 4 | Loading | Stability |
|---|---|---|
| Free.cholesterol.in.large.HDL | 0.97 | 0.4 |
| Total.lipids.in.very.large.HDL | 0.22 | 1 |
| Cholesteryl.esters.in.very.large.HDL | 0.09 | 0.8 |
| Cholesterol.in.very.large.HDL | 0.07 | 0.8 |
| cd300lg | 0.45 | 1 |
| apom | 0.44 | 1 |
| apod | 0.32 | 1 |
| art3 | 0.28 | 1 |
| apoc1 | 0.27 | 1 |
| rgma | 0.27 | 1 |
| pltp | 0.24 | 1 |
| clec4a | 0.22 | 1 |
| angptl3 | 0.21 | 1 |
| apoa1 | 0.17 | 1 |
| notch3 | 0.17 | 0.8 |
| mxra8 | 0.16 | 1 |
| cd248 | 0.13 | 0.8 |
| hrc | 0.12 | 0.8 |
| ca14 | 0.09 | 0.6 |
| adipoq | 0.07 | 0.8 |
| igfbp2 | 0.01 | 0.6 |
| itgb6 | 0.01 | 0.2 |
| tgfbr3 | 0.01 | 0.4 |
| bpifb1 | 0 | 0.6 |

**Table S4**. List of proteomic and metabolomic features selected in DIABLO Component 4. For each feature, the loading indicates its weight in defining the component, while stability reflects the frequency with which the feature was selected across cross-validation folds, providing a measure of robustness.

| Variable | Units | Weight |
|---|---|---|
| Albumin | g/L | -0.0336 |
| Creatinine | umol/L | 0.0095 |
| Glucose | mmol/L | 0.1953 |
| C-reactive protein (log) | mg/dL | 0.0954 |
| Lymphocyte % | % | -0.0120 |
| Mean cell volume | fL | 0.0268 |
| Red cell distribution width | % | 0.3306 |
| Alkaline phosphatase | U/L | 0.0019 |
| White blood cell count | 1000 cells/uL | 0.0554 |
| Age | Years | 0.0804 |
| Constant | | -19.9067 |
| Gamma | | 0.0077 |

**Table S5:** Phenotypic Aging Variables and Gompertz Coefficients including the regression constant and gamma parameter of the Gompertz model from Levine et al. (2018)[5].

| Metabolite | logFC | AveExpr | adj.P.Val | FC |
|---|---|---|---|---|
| Albumin | -0.04 | 3.69 | 6.51E-242 | 1.03 |
| Glucose.lactate | 0.07 | 1.81 | 1.84E-238 | 1.05 |
| Glucose | 0.07 | 1.53 | 1.84E-136 | 1.05 |
| Triglycerides.to.total.lipids.ratio.in.large.LDL | 0.07 | 1.97 | 2.90E-118 | 1.05 |
| Triglycerides.to.total.lipids.ratio.in.medium.LDL | 0.07 | 1.84 | 9.24E-102 | 1.05 |
| Triglycerides.to.total.lipids.ratio.in.IDL | 0.07 | 2.22 | 1.13E-95 | 1.05 |
| Ratio.of.docosahexaenoic.acid.to.total.fatty.acids | -0.06 | 1.07 | 8.28E-79 | 1.04 |
| Triglycerides.to.total.lipids.ratio.in.small.LDL | 0.06 | 1.84 | 1.90E-68 | 1.04 |
| Total.cholesterol | -0.04 | 1.72 | 1.12E-60 | 1.03 |
| Lactate | 0.05 | 1.57 | 1.81E-60 | 1.04 |
| Clinical.LDL.cholesterol | -0.05 | 1.25 | 6.79E-58 | 1.04 |
| Ratio.of.omega.3.fatty.acids.to.total.fatty.acids | -0.06 | 1.65 | 3.37E-57 | 1.05 |
| Triglycerides.to.total.lipids.ratio.in.very.small.VLDL | 0.04 | 3.01 | 3.44E-56 | 1.03 |
| Cholesterol.to.total.lipids.ratio.in.medium.VLDL | -0.06 | 3.40 | 6.80E-56 | 1.04 |
| Cholesteryl.esters.to.total.lipids.ratio.in.medium.VLDL | -0.09 | 2.79 | 3.67E-55 | 1.06 |
| Cholesteryl.esters.to.total.lipids.ratio.in.very.large.VLDL | -0.07 | 2.86 | 8.59E-44 | 1.05 |
| Total.cholesterol.minus.HDL.C | -0.04 | 1.44 | 3.01E-42 | 1.03 |
| Cholesterol.to.total.lipids.ratio.in.very.large.VLDL | -0.05 | 3.39 | 5.51E-42 | 1.03 |
| Triglycerides.to.total.lipids.ratio.in.small.HDL | 0.05 | 1.69 | 9.69E-40 | 1.03 |
| Triglycerides.to.total.lipids.ratio.in.medium.HDL | 0.05 | 1.80 | 2.26E-31 | 1.04 |
| Ratio.of.omega.6.fatty.acids.to.omega.3.fatty.acids | 0.06 | 2.32 | 7.51E-29 | 1.04 |
| Triglycerides.to.total.lipids.ratio.in.large.HDL | 0.06 | 1.79 | 1.57E-23 | 1.04 |
| Triglycerides.to.total.lipids.ratio.in.very.large.HDL | 0.06 | 1.73 | 1.08E-18 | 1.04 |
| Cholesteryl.esters.to.total.lipids.ratio.in.chylomicrons.and.extremely.large.VLDL | -0.04 | 2.81 | 8.22E-13 | 1.03 |
| Phospholipids.to.total.lipids.ratio.in.chylomicrons.and.extremely.large.VLDL | 0.05 | 2.71 | 8.09E-11 | 1.04 |

**Table S6.** Differential analysis of metabolites between *Accelerated* and *NotAccelerated* aging groups. Results are from limma linear models adjusted for age and sex, with Benjamini–Hochberg FDR correction. Features shown here passed adjusted p-value < 0.05 and fold-change thresholds. Columns show log2 fold-change (logFC), average expression (AveExpr), adjusted p-value (adj.P.Val), and back-transformed fold change (FC). Positive logFC values indicate higher levels in *Accelerated* individuals, and negative values indicate higher levels in *NotAccelerated* individuals.

| Protein | logFC | AveExpr | adj.P.Val | FC |
|---|---|---|---|---|
| clec4d | 0.40 | 0.00 | 0 | 1.32 |
| fstl3 | 0.21 | 0.02 | 0 | 1.15 |
| hgf | 0.27 | 0.02 | 0 | 1.20 |
| igfbp4 | 0.27 | 0.02 | 0 | 1.20 |
| il1rn | 0.38 | 0.08 | 0 | 1.30 |
| il6 | 0.49 | 0.11 | 0 | 1.40 |
| lilra5 | 0.21 | 0.00 | 0 | 1.15 |
| osm | 0.55 | 0.00 | 0 | 1.47 |
| plaur | 0.21 | 0.01 | 0 | 1.15 |
| tgfa | 0.35 | 0.07 | 0 | 1.27 |
| tnfrsf1a | 0.21 | 0.02 | 0 | 1.16 |
| vsig4 | 0.26 | 0.02 | 0 | 1.20 |
| gdf15 | 0.27 | 0.04 | 0.00E+00 | 1.21 |
| mmp9 | 0.37 | -0.02 | 5.17E-303 | 1.30 |
| cd300e | 0.25 | 0.03 | 1.00E-302 | 1.19 |
| wfdc2 | 0.22 | 0.05 | 1.65E-281 | 1.16 |
| ceacam8 | 0.28 | -0.01 | 3.20E-280 | 1.21 |
| col6a3 | 0.20 | 0.01 | 6.75E-277 | 1.15 |
| tnfrsf1b | 0.20 | 0.03 | 1.08E-276 | 1.15 |
| angptl4 | 0.26 | -0.01 | 1.48E-274 | 1.20 |
| prap1 | 0.25 | -0.03 | 1.54E-274 | 1.19 |
| lcn2 | 0.21 | 0.01 | 7.51E-269 | 1.15 |
| inhbb | 0.26 | 0.02 | 1.92E-247 | 1.20 |
| fabp4 | 0.34 | 0.03 | 6.95E-244 | 1.26 |
| rarres2 | 0.28 | 0.00 | 4.49E-242 | 1.21 |
| chchd10 | 0.20 | 0.02 | 9.49E-241 | 1.15 |
| retn | 0.22 | 0.01 | 4.05E-239 | 1.17 |
| mmp8 | 0.39 | 0.02 | 2.58E-230 | 1.31 |
| tnfrsf11a | 0.20 | 0.01 | 9.17E-227 | 1.15 |
| lair1 | 0.20 | 0.02 | 1.86E-225 | 1.15 |
| fgf23 | 0.28 | 0.08 | 4.28E-214 | 1.22 |
| pglyrp1 | 0.22 | 0.01 | 2.21E-209 | 1.16 |
| mzb1 | 0.26 | 0.02 | 7.64E-200 | 1.20 |
| olr1 | 0.28 | 0.04 | 7.37E-187 | 1.21 |
| lep | 0.47 | -0.08 | 6.08E-184 | 1.38 |
| clec6a | 0.22 | 0.01 | 1.47E-183 | 1.16 |
| lbp | 0.27 | -0.02 | 1.64E-177 | 1.21 |
| fabp3 | 0.22 | 0.05 | 1.37E-174 | 1.17 |
| fcn1 | 0.24 | -0.03 | 6.26E-172 | 1.18 |
| prtn3 | 0.20 | 0.01 | 4.88E-169 | 1.15 |
| areg | 0.20 | 0.06 | 5.31E-165 | 1.15 |

| | | | | |
|---|---|---|---|---|
| defa1_defa1b | 0.22 | 0.03 | 1.00E-158 | 1.16 |
| rbp7 | 0.25 | 0.06 | 3.58E-152 | 1.19 |
| tnfrsf6b | 0.24 | 0.04 | 4.82E-146 | 1.18 |
| prg2 | 0.20 | 0.03 | 4.94E-146 | 1.15 |
| vegfa | 0.24 | 0.08 | 2.40E-138 | 1.18 |
| lamp3 | 0.23 | 0.05 | 6.66E-134 | 1.17 |
| sele | 0.22 | -0.02 | 6.98E-134 | 1.16 |
| fgl1 | 0.23 | -0.01 | 6.55E-128 | 1.18 |
| serpinb8 | 0.22 | 0.01 | 5.01E-126 | 1.16 |
| dpy30 | 0.24 | 0.03 | 2.52E-125 | 1.18 |
| bpifb2 | 0.25 | 0.13 | 2.90E-124 | 1.19 |
| s100a12 | 0.29 | -0.02 | 4.82E-124 | 1.22 |
| pi3 | 0.21 | 0.03 | 2.26E-123 | 1.16 |
| cst7 | 0.37 | 0.12 | 6.49E-122 | 1.29 |
| chi3l1 | 0.30 | 0.11 | 3.25E-121 | 1.23 |
| trem2 | 0.20 | 0.03 | 8.29E-120 | 1.15 |
| ccl7 | 0.23 | 0.05 | 4.92E-119 | 1.17 |
| ccl3 | 0.21 | 0.05 | 1.41E-113 | 1.16 |
| ccl20 | 0.33 | 0.13 | 2.68E-113 | 1.26 |
| tnfsf14 | 0.22 | 0.01 | 8.29E-113 | 1.16 |
| baiap2 | 0.24 | 0.06 | 5.62E-111 | 1.18 |
| fcamr | 0.26 | 0.03 | 2.40E-106 | 1.20 |
| il19 | 0.24 | 0.07 | 4.60E-106 | 1.18 |
| igsf9 | 0.31 | 0.08 | 1.52E-103 | 1.24 |
| tff1 | 0.28 | 0.15 | 2.21E-100 | 1.21 |
| capg | 0.21 | -0.03 | 9.20E-100 | 1.15 |
| gast | 0.42 | 0.38 | 1.29E-99 | 1.34 |
| havcr1 | 0.22 | 0.03 | 1.36E-99 | 1.16 |
| gusb | 0.22 | 0.04 | 2.69E-98 | 1.17 |
| cd300lf | 0.21 | -0.13 | 7.53E-98 | 1.16 |
| tff2 | 0.20 | 0.05 | 2.01E-97 | 1.15 |
| rbp5 | 0.21 | 0.07 | 1.47E-95 | 1.15 |
| ncf2 | 0.28 | 0.04 | 1.56E-94 | 1.22 |
| calca | 0.21 | 0.11 | 2.78E-94 | 1.16 |
| grpel1 | 0.21 | 0.07 | 5.96E-94 | 1.15 |
| epo | 0.26 | 0.09 | 1.78E-92 | 1.19 |
| gpr37 | 0.21 | -0.01 | 3.27E-91 | 1.16 |
| tmsb10 | 0.26 | 0.00 | 5.29E-90 | 1.20 |
| lbr | 0.22 | 0.05 | 1.04E-87 | 1.17 |
| krt18 | 0.28 | 0.12 | 6.43E-85 | 1.21 |
| fabp1 | 0.31 | 0.05 | 2.97E-82 | 1.24 |
| azu1 | 0.23 | 0.07 | 5.19E-82 | 1.18 |

| | | | | |
|---|---|---|---|---|
| sncg | 0.22 | -0.04 | 6.00E-82 | 1.16 |
| chga | 0.25 | 0.13 | 2.57E-80 | 1.19 |
| ren | 0.23 | 0.09 | 5.68E-80 | 1.17 |
| bgn | -0.36 | 0.13 | 6.57E-79 | 1.28 |
| il17c | 0.21 | 0.09 | 8.97E-78 | 1.16 |
| olfm4 | 0.33 | -0.10 | 8.07E-75 | 1.25 |
| mnda | 0.29 | 0.04 | 4.71E-72 | 1.22 |
| crh | -0.25 | 0.05 | 1.65E-69 | 1.19 |
| ccl18 | 0.21 | 0.04 | 7.13E-69 | 1.15 |
| rnase3 | 0.31 | 0.06 | 2.63E-68 | 1.24 |
| mvk | 0.23 | 0.05 | 3.36E-67 | 1.17 |
| hpse | 0.25 | -0.03 | 2.11E-65 | 1.19 |
| alpp | 0.35 | 0.13 | 3.42E-64 | 1.27 |
| fgf21 | 0.35 | 0.05 | 7.11E-62 | 1.27 |
| aifm1 | 0.29 | 0.07 | 1.61E-61 | 1.22 |
| pyy | 0.25 | 0.10 | 3.19E-61 | 1.19 |
| cxcl11 | 0.21 | 0.06 | 2.47E-60 | 1.16 |
| mmp1 | 0.24 | -0.05 | 5.23E-58 | 1.18 |
| ca5a | 0.25 | 0.09 | 1.07E-57 | 1.19 |
| echs1 | 0.22 | 0.04 | 1.17E-51 | 1.17 |
| insl5 | 0.23 | 0.14 | 1.55E-51 | 1.17 |
| ccl17 | 0.23 | 0.00 | 1.55E-51 | 1.17 |
| oxt | 0.38 | 0.14 | 5.45E-51 | 1.30 |
| dut | 0.23 | 0.01 | 1.47E-47 | 1.18 |
| defb4a_defb4b | 0.33 | 0.04 | 3.30E-43 | 1.26 |
| upb1 | 0.20 | 0.09 | 2.96E-40 | 1.15 |
| eif4g1 | 0.21 | -0.09 | 3.83E-35 | 1.16 |
| ccl5 | 0.21 | 0.00 | 7.12E-35 | 1.16 |
| bpifa2 | -0.20 | 0.05 | 1.89E-34 | 1.15 |
| gcg | 0.25 | 0.05 | 7.98E-33 | 1.19 |
| timp3 | 0.26 | 0.16 | 2.05E-31 | 1.19 |
| paep | 0.26 | 0.55 | 2.28E-31 | 1.19 |
| hao1 | 0.24 | 0.15 | 2.09E-26 | 1.18 |
| siglec5 | 0.20 | -0.31 | 1.70E-24 | 1.15 |
| yars1 | 0.22 | -0.16 | 2.86E-21 | 1.16 |
| gh1 | -0.22 | 0.20 | 1.18E-12 | 1.16 |

**Table S7.** Differential analysis of proteins between *Accelerated* and *NotAccelerated* aging groups. Results are from limma linear models adjusted for age and sex, with Benjamini–Hochberg FDR correction. Features shown here passed adjusted p-value < 0.05 and fold-change thresholds. Columns show log2 fold-change (logFC), average expression (AveExpr), adjusted p-value (adj.P.Val), and back-transformed fold change (FC). Positive logFC values indicate higher levels in *Accelerated* individuals, and negative values indicate higher levels in *NotAccelerated* individuals.

| Genes | Fibro IR | Fibro RAS | Fibro ATV | Epi IR | Exosome IR | Exosome RAS |
|---|---|---|---|---|---|---|
| **APCS** | | | | | 2.28 | |
| **APOA1** | | 1.7 | | | 2.83 | |
| **APOD** | | | 0.98 | | | |
| **APOM** | | | | | -2.08 | |
| **AXL** | | | | -2.52 | | |
| **B4GAT1** | | | | -0.55 | | |
| **COL6A3** | 0.48 | 1.5 | 2.06 | | -2.5 | -2.78 |
| **DPP4** | 7.83 | | | 0.35 | | |
| **GDF15** | 2.54 | 5.07 | 3.08 | 1.96 | | |
| **IGFBP2** | 5.82 | 2.61 | 0.42 | 1.24 | | |
| **IGFBP3** | 3.79 | 0.68 | 0.53 | 0.17 | | |
| **IGFBP4** | 3.25 | 1.71 | 2.1 | -1.86 | | 3.5 |
| **LAMP2** | -1.54 | | | | | |
| **LCAT** | | | | | -1.7 | |
| **LDLR** | | 2.73 | | -0.73 | | |
| **MASP1** | | | 1.64 | | | |
| **MMP9** | | 6.57 | | -2.07 | | |
| **PLAUR** | | 4.37 | | | | |
| **PLTP** | | 1.36 | 2 | | | |
| **RARRES2** | | | | -2.67 | | |
| **RSE4** | | 2.72 | 1.86 | | | |
| **SPOCK1** | | 1.4 | 2.62 | -1.28 | | |
| **TFRC** | | 4.64 | | | | |
| **VCAM1** | | | | -1.09 | | |

**Table S8. $Log_2$ fold-change values of DIABLO-selected proteins across SASP Atlas senescence models.** The table lists the $log_2$ fold-change (senescent vs. control) protein abundance values extracted from the SASP Atlas, corresponding to the heatmap shown in **Supplementary Figure S8**. Values are shown for multiple senescence models: fibroblast irradiation (Fibro IR), fibroblast RAS activation (Fibro RAS), fibroblast atazanavir-induced (Fibro ATV), epithelial irradiation (Epi IR), and exosome fractions from irradiated and RAS-induced cells (Exosome IR, Exosome RAS). Positive values indicate upregulation in senescent cells, and negative values indicate downregulation. Missing values represent proteins not detected or not significantly changed in the corresponding SASP Atlas experiment.

| Full metabolite name | Abbreviation used in plots |
|---|---|
| Total lipids in lipoprotein particles | TL.lp |
| Polyunsaturated fatty acids | PUFAs |
| Ratio of monounsaturated fatty acids to total fatty acids | MUFA/TFA |
| Glycoprotein acetyls | GlycA |
| Total lipids in medium VLDL | TL.M_VLDL |
| Cholesteryl esters in medium LDL | CE.M_LDL |
| Glucose:lactate ratio | Glu/Lact |
| Glutamine | Glutamine |
| Total lipids in very large HDL | TL.VL_HDL |
| Cholesterol in very large HDL | Chol.VL_HDL |
| Cholesteryl esters in very large HDL | CE.VL_HDL |
| Cholesteryl esters in very small VLDL | CE.VS_VLDL |
| Free cholesterol in large HDL | F.Chol.L_HDL |

**Table S9. Metabolite abbreviations used in figures.** For clarity, long metabolite names were shortened to abbreviated forms in figures (loadings, stability plots, and network representations). Full names correspond to variables measured in the Nightingale NMR metabolomics platform of the UK Biobank.
